# Does Education Sculpt Healthcare Choices? Exploring Factors Influencing Healthcare Utilization Among Female Youths in Eight Low and Lower-Middle-Income Countries

**DOI:** 10.1101/2024.02.01.24302136

**Authors:** Nahid Hassan Nishan, Khadiza Akter, Afroza Sharmin, Tazmin Akter Tithi, M Z E M Naser Uddin Ahmed

## Abstract

**Introduction:** Emphasizing the global commitment to universal health coverage, this research addresses geographical variations and challenges faced by young females across low and lower-middle-income countries. Therefore, the objective of this study is to determine the factors that influence the decision-making of young females when it comes to seeking healthcare services in low or lower-middle-income countries.

**Methodology:** We examined healthcare usage among female youth across eight countries. We used data from the DHS and employed regression and Chi^2^ tests for our analysis. Our focus was on females aged 15 to 24 and their visits to healthcare facilities. To ensure the validity of our findings, we used weighted sampling to represent the population.

**Results:** We had a total sample of 51,298 female youth groups between the ages of 15 and 24 who participated in our study. When it comes to the demographics of the participants, most of those in the 15-19 age group were from Burkina Faso (54.26%), while most of those in the 20-24 age group were from Ghana (50.19%). The impact of education varied across countries; primary education led to increased healthcare utilization in Kenya, Tanzania, and Cambodia, but unexpected trends were observed in Burkina Faso.

**Conclusions:** Education significantly influences healthcare utilization, both primary and secondary education positively impacting this. Rural residents face challenges accessing timely healthcare. Geographical challenges, like diseases and limited accessibility, contribute to varied healthcare usage in Kenya and the Philippines. Addressing infrastructure issues is critical, emphasizing education and promoting transparency to enhance healthcare equity.

## Introduction

Healthcare utilization encompasses the degree to which people use the services of the healthcare system [1]. Several factors determine this healthcare utilization, and it can be broadly categorized into two types: service-related and user-related factors. Service factor refers to the accessibility, availability, and quality of the service. Conversely, user factors include individual characteristics such as age, gender, other demographics, specific health needs, and perceived beliefs regarding their health and the healthcare system [1,2]. Though several factors determine healthcare utilization, the ultimate goal of this healthcare utilization is to protect and promote people’s health [1]. More importantly, there is a global desire to ensure healthcare utilization for individuals irrespective of their barriers. This commitment is a crucial target that has been declared in the 2030 Agenda [3]. Specifically, this falls under the Sustainable Development Goal (SDG), and the number three goal emphasizes the achievement of universal health coverage to enhance health promotion and well-being for people of all ages, irrespective of their demographic characteristics [4]. So, different organizations (for example, UNDP) and governments are working progressively to achieve this goal [5]. It is believed that the functional healthcare system is very crucial to reaching the SDG goal and supporting universal health coverage by ensuring healthcare utilization [6]. However, evidence shows that the rate of healthcare utilization varies due to geographical differences, and this rate could mostly vary in low and lower-middle-income countries across different population groups [7–9]. Their financial hardship and the underdeveloped healthcare system pose a significant barrier [6]. Considering this issue, global experts are giving special attention to low and lower-middle-income countries so that they can have better healthcare utilization and ensure quality healthcare [10].

As this progressed, studies showed that the majority focused on adult age group people, children under five years old, and often the elderly. For this, there are several incorporated activities in low and lower-middle-income countries, such as insurance coverage for maternal healthcare utilization and primary healthcare services are introduced [11–13]. Even though there have been improvements over the three decades, children and teenagers in low-income and middle-income countries are still not achieving their full health potential [14]. This is more intense, in low and lower-middle-income countries as this creates burden in many aspect of utilization of healthcare among young female groups [15]. It is crucial to focus on them as studies have shown they are vulnerable to several adverse health consequences, such as poor reproductive health, chronic diseases, complications from substance abuse, mental health disorders, behavioral complexity, suicidal ideation, and many more [14,16,17]. Additionally, these groups face tremendous challenges in seeking healthcare services, and often, in several cases, it is difficult for the female youth as they face more challenges than adult females while receiving healthcare services [18]. Youth groups in lower-middle-income countries face a range of health challenges that affect different aspects of their well-being. Accidental injuries and road traffic accidents are a cause of death and disability among them. Infectious diseases like HIV, tuberculosis, and respiratory infections still impact adolescents, while the ongoing COVID-19 pandemic adds difficulties to their well-being [19]. Early pregnancy and childbirth environmental risks, overweight issues, and malnutrition add to the complexity of health problems faced by this group [19]. In addition, the long distance of healthcare makes it difficult for young females to receive proper healthcare services [20]. So, it can be seen that there are multiple factors that influence the reception of healthcare services among youth groups, and this may not be the same across low- and lower-middle-income countries. This raises the question of which factors mostly play a vital role in utilizing health care among young female groups in those countries. If we can identify those crucial factors that are mostly contributing to healthcare utilization among low and lower-middle-income countries, this finding will help the stakeholders and policymakers to consider more effective implementation and intervention to reach the SDG goal properly, and this poses a great novelty in this study. After through literature review, we identified this research gap and therefore, this study aims to identify the factors influencing the female youth’s decision to receive healthcare services in low- and lower-middle-income countries.

## Method and Materials

We relied on the secondary dataset from the Demographic and Health Survey (DHS) to conduct this study. The DHS contains data from 108 countries globally [21]. However, we chose 62 countries categorized as low-income and lower-middle-income countries by the World Bank ranking system [22]. Eight of these nations have been chosen because they met our interest criterion: the utilization of healthcare facilities among youth groups. The countries enlisted for this study are Cross-sectional study techniques have been followed to complete this study using the data extracted from the most updated DHS dataset from above 2020 though we started searching the dataset from 2017 but none of them contain all the variables suitable for our study. This DHS survey usually follows a two-step sampling approach to get the data from a desired country. Firstly, it selects the household in-cluster enumeration areas (EAs), and then a defined number of households are chosen from each cluster. The DHS collaborates and empowers the national implementing agencies through regional workshops so that they can ensure the training and other necessary requirements for the field staff to collect data effectively from the household. For detailed information regarding their study site, population, data collection, and sampling process, you may visit [31].

For this study, we considered the individual recorded information (IR files) obtained from the DHS as it has access to data of females aged 15 to 49 years. Our goal was to get information regarding whether the young generation (female) has utilized health facilities in recent years, so we took the variable “visited a health facility in the last 12 months”. We restricted it to those above 24 years. We also ensure that there is no presence of missing values. Moreover, any unwanted variables have also been removed. Additionally, weight has been applied to the sample of all countries to adjust them according to the population sizes and survey years.

### Dependent and Independent Variables

The outcome variable we have used for this study is whether the respondent visited a healthcare facility in the last twelve months. Depending upon several condition there could be many reasons an individual will visit the health care settings [32]. However, for this current study, we only considered a particular group (youth) who received healthcare services through healthcare visit. Moreover, we kept the outcome variable in a binary approach with values 1 and 0 for analysis based on the study objectives. Here, value “1” refers to visited healthcare facilities in the last twelve months, and “0” refers otherwise.

Initially, a total of 11 variables were incorporated as independent variables for this current study due to their access to DHS in these seven countries. However, within these 11 variables, we have modified it to seven explanatory variables to align with our research interest. These variables encompass age, residence, wealth index, personal access to vehicle, reaching time to nearest health care facility, and mass media engagement. The primary explanatory variable is the level of education. For our analysis, we have categorized these variables in a specific way. For example, an ordinal scale has been employed for the age variable: “15-19 Age group” and “20-24 Age group”. The education variable is categorized as individual has No education, Primary, Secondary, and Higher. The resident has kept a dichotomy: Urban and Rural.

Furthermore, the wealth index is categorized as Poor, Middle, and Rich. Personal access to a vehicle is determined by combining three variables (household having a bicycle, household having motorcycle/scooter, household having car/truck) and then categorizing them as “Yes”-have access and “No” have no access. Additionally, reaching time to the nearest healthcare facility has been categorized as “Within 1 hour”, “2-3 Hours,” and “More than 3 hours”. Finally, mass media engagement has been categorized as “Present” media engagement” and “Absent” media engagement by merging three variables (frequency of reading newspapers or magazines, frequency of watching TV, and frequency of listening to radio).

### Data analysis

To analyze our data thoroughly, we relied on Stata, a statistical software version 17. As DHS has a two-stage cluster sampling design, there is a potential for unequal sampling probability, clustering, and stratification. However, we applied the weighting technique through survey (Svy) commands to resolve this issue. The weighting technique enables the fitting of a proper statistical model by ensuring the representativeness of the survey data, and DHS highly recommends this technique [33].

We begin our analysis by calculating descriptive statistics to present a comprehensive overview of the variables. Afterward, we did the bivariate analysis. We checked the association through the Chi^2^ test, which helped us to assess the explanatory variables’ significance level. We used the (Ψ) symbol to highlight the Chi^2^ significant strengths. We individually examined all explanatory variables to assess their association with the outcome variable and found that all of the independent variables we chose were suitable for multivariate analysis as their *P*-value were < 0.05. Moving forward to multivariate analysis, we applied logistic regression, carefully observed the *P*-value, and presented the significance with the adjusted odd ratio (AOR). To show multivariate AOR strength, we used (*), where more stars represent more strength. We considered a 95% confidence interval for this multivariate analysis. In addition, with these statistical analyses, we have also checked multicollinearity and confirmed there was no presence of multicollinearity, and the model was fitted properly for our analysis. Lastly, we constructed tables to present our study findings.

## Results

We got a total of 51,298 weighted youth aged 15 to 24 participants in our study. In terms of demographic characteristics, most participant’s 15-19-year age group were in Burkina Faso (54.26%), and most participants from the 20-24 years group were from Ghana (50.19%). The majority of the participants from Nepal live in urban areas (68.09%), and most Burkina Faso participants are from rural areas (64.84%). About (43.90%) received primary education, whereas (68.86%) and (27.90%) received secondary and higher education in the Philippines. More information is in **Table 2**

**Table 1:**
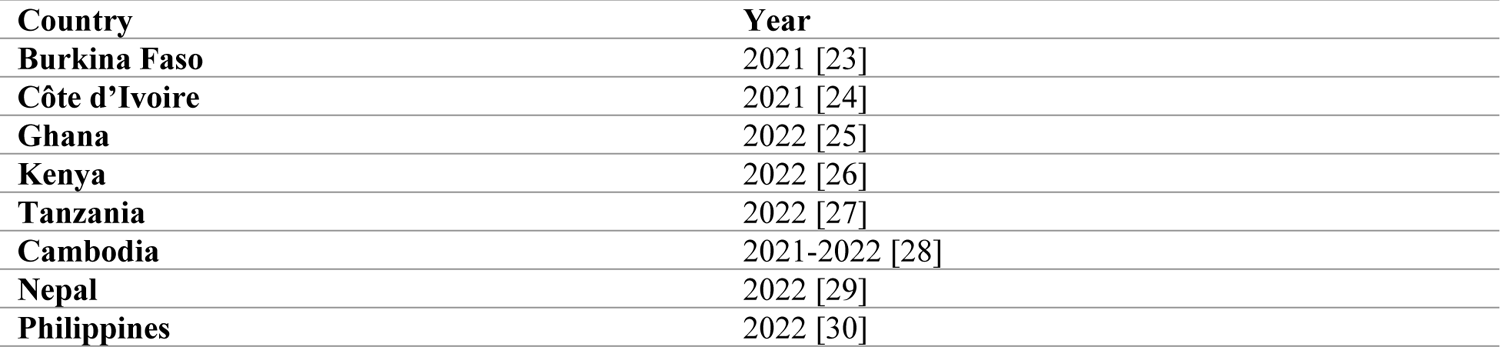
List of Countries Used In this Study.

| Country | Year |
| --- | --- |
| Burkina Faso | 2021 [23] |
| Côte d'Ivoire | 2021 [24] |
| Ghana | 2022 [25] |
| Kenya | 2022 [26] |
| Tanzania | 2022 [27] |
| Cambodia | 2021-2022 [28] |
| Nepal | 2022 [29] |
| Philippines | 2022 [30] |

**Table 2:**
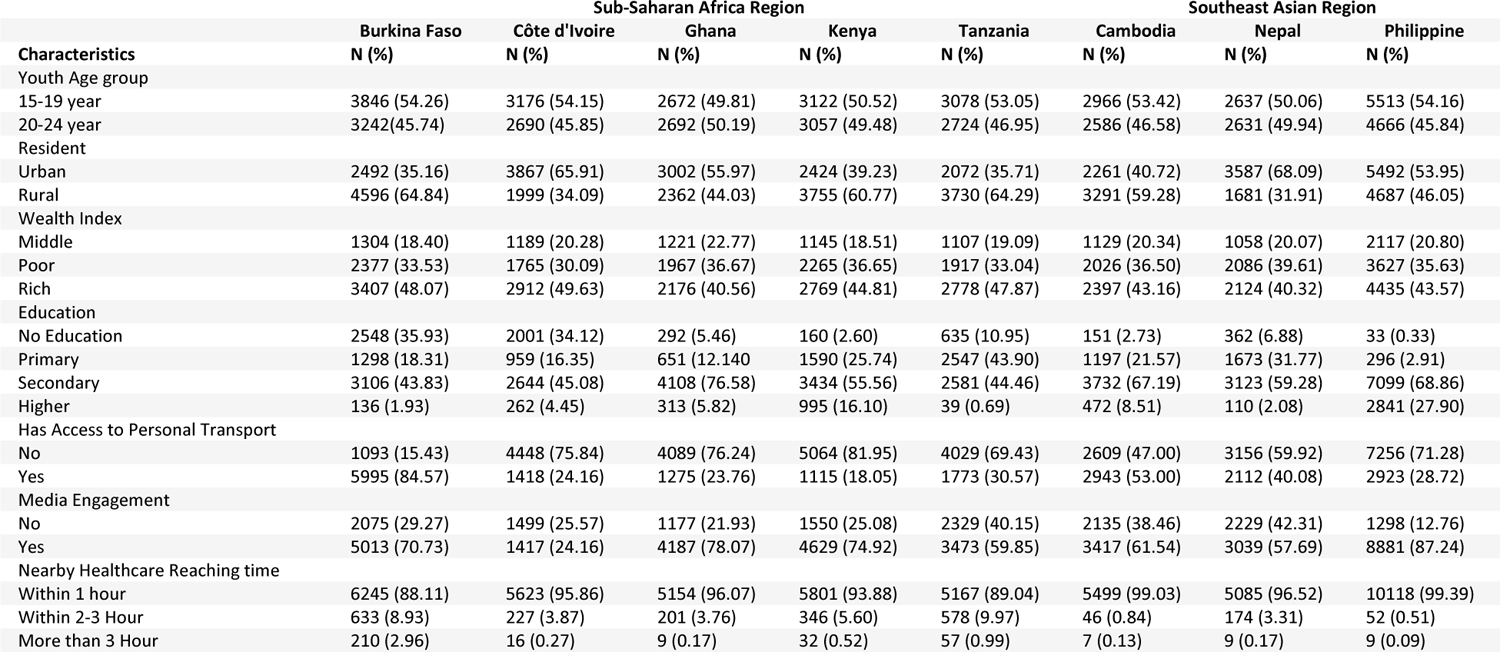
Demographic Characteristics of Individuals.

| Characteristics | Sub-Saharan Africa Region |  |  |  |  | Southeast Asian Region |  |  |
| --- | --- | --- | --- | --- | --- | --- | --- | --- |
|  | Burkina Faso<br>N (%) | Côte d'Ivoire<br>N (%) | Ghana<br>N (%) | Kenya<br>N (%) | Tanzania<br>N (%) | Cambodia<br>N (%) | Nepal<br>N (%) | Philippine<br>N (%) |
| Youth Age group |  |  |  |  |  |  |  |  |
| 15-19 year | 3846 (54.26) | 3176 (54.15) | 2672 (49.81) | 3122 (50.52) | 3078 (53.05) | 2966 (53.42) | 2637 (50.06) | 5513 (54.16) |
| 20-24 year | 3242(45.74) | 2690 (45.85) | 2692 (50.19) | 3057 (49.48) | 2724 (46.95) | 2586 (46.58) | 2631 (49.94) | 4666 (45.84) |
| Resident |  |  |  |  |  |  |  |  |
| Urban | 2492 (35.16) | 3867 (65.91) | 3002 (55.97) | 2424 (39.23) | 2072 (35.71) | 2261 (40.72) | 3587 (68.09) | 5492 (53.95) |
| Rural | 4596 (64.84) | 1999 (34.09) | 2362 (44.03) | 3755 (60.77) | 3730 (64.29) | 3291 (59.28) | 1681 (31.91) | 4687 (46.05) |
| Wealth Index |  |  |  |  |  |  |  |  |
| Middle | 1304 (18.40) | 1189 (20.28) | 1221 (22.77) | 1145 (18.51) | 1107 (19.09) | 1129 (20.34) | 1058 (20.07) | 2117 (20.80) |
| Poor | 2377 (33.53) | 1765 (30.09) | 1967 (36.67) | 2265 (36.65) | 1917 (33.04) | 2026 (36.50) | 2086 (39.61) | 3627 (35.63) |
| Rich | 3407 (48.07) | 2912 (49.63) | 2176 (40.56) | 2769 (44.81) | 2778 (47.87) | 2397 (43.16) | 2124 (40.32) | 4435 (43.57) |
| Education |  |  |  |  |  |  |  |  |
| No Education | 2548 (35.93) | 2001 (34.12) | 292 (5.46) | 160 (2.60) | 635 (10.95) | 151 (2.73) | 362 (6.88) | 33 (0.33) |
| Primary | 1298 (18.31) | 959 (16.35) | 651 (12.140) | 1590 (25.74) | 2547 (43.90) | 1197 (21.57) | 1673 (31.77) | 296 (2.91) |
| Secondary | 3106 (43.83) | 2644 (45.08) | 4108 (76.58) | 3434 (55.56) | 2581 (44.46) | 3732 (67.19) | 3123 (59.28) | 7099 (68.86) |
| Higher | 136 (1.93) | 262 (4.45) | 313 (5.82) | 995 (16.10) | 39 (0.69) | 472 (8.51) | 110 (2.08) | 2841 (27.90) |
| Has Access to Personal Transport |  |  |  |  |  |  |  |  |
| No | 1093 (15.43) | 4448 (75.84) | 4089 (76.24) | 5064 (81.95) | 4029 (69.43) | 2609 (47.00) | 3156 (59.92) | 7256 (71.28) |
| Yes | 5995 (84.57) | 1418 (24.16) | 1275 (23.76) | 1115 (18.05) | 1773 (30.57) | 2943 (53.00) | 2112 (40.08) | 2923 (28.72) |
| Media Engagement |  |  |  |  |  |  |  |  |
| No | 2075 (29.27) | 1499 (25.57) | 1177 (21.93) | 1550 (25.08) | 2329 (40.15) | 2135 (38.46) | 2229 (42.31) | 1298 (12.76) |
| Yes | 5013 (70.73) | 1417 (24.16) | 4187 (78.07) | 4629 (74.92) | 3473 (59.85) | 3417 (61.54) | 3039 (57.69) | 8881 (87.24) |
| Nearby Healthcare Reaching time |  |  |  |  |  |  |  |  |
| Within 1 hour | 6245 (88.11) | 5623 (95.86) | 5154 (96.07) | 5801 (93.88) | 5167 (89.04) | 5499 (99.03) | 5085 (96.52) | 10118 (99.39) |
| Within 2-3 Hour | 633 (8.93) | 227 (3.87) | 201 (3.76) | 346 (5.60) | 578 (9.97) | 46 (0.84) | 174 (3.31) | 52 (0.51) |
| More than 3 Hour | 210 (2.96) | 16 (0.27) | 9 (0.17) | 32 (0.52) | 57 (0.99) | 7 (0.13) | 9 (0.17) | 9 (0.09) |

In a bivariate analysis, there is a highly significant association (*p-*value <0.001) present between the age group and visiting healthcare to receive healthcare facilities, and this significance is present in all of the countries of our study. Moreover, the residents have a strong association with outcomes in Cambodia and the Philippines (*p-*value <0.001), but Côte d’Ivoire showed a weak statistical association (*p-*value <0.05). Furthermore, education showed a high statistically significant association (*p-*value <0.001) in multiple countries (Burkina Faso, Côte d’Ivoire, Ghana, Kenya, Cambodia, and the Philippines). Additionally, the wealth index and media engagement also showed a high statistically significant association (*p*-value <0.001) in Kenya, Cambodia, and the Philippines, but a moderate association (*p*-value <0.01) was only found for the wealth index and a weak association (*p-*value <0.05) for media engagement in Burkina Faso. Other variables include access to personal transport, having a moderate association (*p*-value <0.01) in Tanzania and Nepal, and reaching time to nearby healthcare facilities, which only showed a weak association (*p-*value <0.05) in Nepal.

### Demographic Factor

In our study, age plays a significant role in healthcare utilization among youth. Individuals aged 20-24 years across Burkina Faso, Côte d’Ivoire, Ghana, Kenya, Tanzania, Cambodia, Nepal, and the Philippines showed a higher likelihood of accessing healthcare services compared to the 15-19 years age group. The adjusted odds ratios are respectively: (AOR: 3.44 CI:3.04-3.89), (AOR: 2.75 CI: 2.39-3.17), (AOR: 2.41 CI: 2.08-2.76), (AOR: 2.46 CI: 2.11-2.87), (AOR: 2.93, CI: 2.55-3.37), (AOR: 2.25, CI: 1.91-2.64), (AOR: 2.64, CI: 2.32-3.01), and (AOR: 3.87, CI: 3.22-4.64) and all of them are highly statistically significant.

The important key factor, education, and its influence on healthcare utilization are evident in Burkina Faso, Côte d’Ivoire, Kenya, Tanzania, and Cambodia. In Kenya, Tanzania, and Cambodia, individuals with primary education showed a higher likelihood (AOR 2.07 CI:1.50-2.85), (AOR 1.36 CI:1.09-1.68), and (AOR 1.55 CI:1.01-2.37) of visiting healthcare facilities compared to those without education except Burkina Faso where there is a lower likelihood in primary education (AOR 0.84 CI:0.72-0.98) compared to those without education, with statistical significance. Additionally, Burkina Faso also showed a lower likelihood (AOR 0.83 CI:0.73-0.95) of secondary education compared to those who are not educated, but Côte d’Ivoire, Kenya, and Tanzania showed the opposite, where they showed a higher likelihood of healthcare utilization (AOR 1.31 CI:1.07-1.57), (AOR 2.58 CI:1.88-3.54) and (AOR 1.42 CI:1.11-1.80), respectively. Only Kenya showed a higher likelihood of (AOR 2.16 CI:1.48-3.17) healthcare utilization among higher educated individuals compared to those who were not educated, and this is also highly significant.

However, in terms of residents, there is an increased likelihood of healthcare utilization seen in rural individuals in the Sub-Saharan African region: Kenya and in Southeast Asian region: Philippines with (AOR 1.41 CI: 1.17-1.70) and (AOR 1.24 CI: 1.00-1.53) compared to the urban people.

### Socio-economic factor

In the wealth index context, Burkina Faso and Cambodia showed a significant role. In Burkina Faso, the poor have a lower likelihood (AOR 0.73 CI:0.62-0.87), and in Cambodia rich also have a lower likelihood (AOR 0.73 CI:0.59-0.92) of healthcare utilization compared to the middle class, and both are significant. In Tanzania and Cambodia, those who have personal transport both have a lower likelihood (AOR 0.84 CI:0.72-0.99) and (AOR 0.71 CI:0.59-0.82) of healthcare utilization compared to those who do not, and this result is also statistically significant.

### Healthcare accessibility

**Table 4** shows a significant association across all the regions of our study between Urban-rural settings and Nearby healthcare reaching time. There is a noticeable trend across all of the regions, which is that there is a higher percentage of individuals who live in urban regions have reached within 1 hour compared to those who live in rural areas.

**Table 3:**
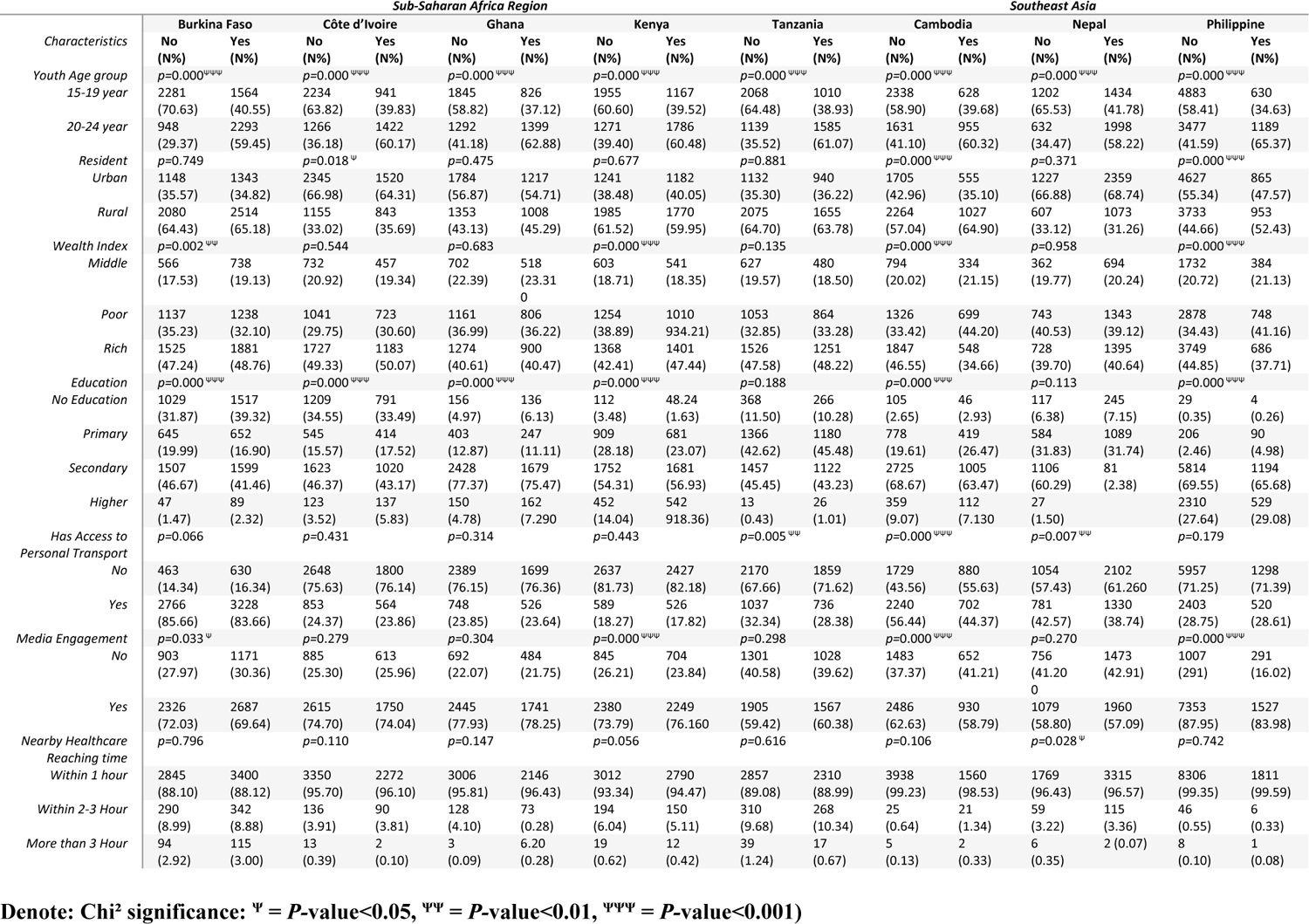
Association between Influencing Factors and Healthcare Utilization.

**Table 4:** Association between Residence and Time Required to Reach Nearby Healthcare Facilities.

| REACHING TIME<br>NEAREST HEALTHCARE | SUB-SAHARAN AFRICA REGION |  |  |  |  |  |  |  | SOUTHEAST ASIA |  |  |  |  |  |  |  |
| --- | --- | --- | --- | --- | --- | --- | --- | --- | --- | --- | --- | --- | --- | --- | --- | --- |
|  | Burkina Faso<br>*p*=0.000 <sup>***</sup> |  | Côte d'Ivoire<br>*p*=0.000 <sup>***</sup> |  | Ghana<br>*p*=0.000 <sup>***</sup> |  | Kenya<br>*p*=0.000 <sup>***</sup> |  | Tanzania<br>*p*=0.000 <sup>***</sup> |  | Cambodia<br>*p*=0.000 <sup>***</sup> |  | Nepal<br>*p*=0.000 <sup>***</sup> |  | Philippine<br>*p*=0.002 <sup>**</sup> |  |
|  | Urban | Rural | Urban | Rural | Urban | Rural | Urban | Rural | Urban | Rural | Urban | Rural | Urban | Rural | Urban | Rural |
| **1 HOUR** | N (%)<br>2424<br>(97.29) | N (%)<br>3820<br>(83.13) | N (%)<br>3776<br>(97.69) | N (%)<br>1846<br>(92.34) | N (%)<br>2954<br>(98.40) | N (%)<br>2199<br>(93.11) | N (%)<br>2370<br>(97.76) | N (%)<br>3432<br>(91.38) | N (%)<br>2021<br>(97.53) | N (%)<br>3146<br>(84.32) | N (%)<br>2256<br>(99.81) | N (%)<br>3242<br>(98.50) | N (%)<br>3501<br>(97.62) | N (%)<br>1583<br>(94.17) | N (%)<br>5471<br>(99.62) | N (%)<br>4646<br>(99.13) |
| **2-3 HOURS** | 22<br>(0.91) | 610<br>(13.28) | 81<br>(2.10) | 145<br>(7.29) | 46<br>(1.53) | 155<br>(6.59) | 53<br>(2.21) | 292<br>(7.78) | 46<br>(2.22) | 532<br>(14.28) | 2<br>(0.08) | 44<br>(1.36) | 81<br>(2.28) | 92<br>(5.53) | 15<br>(0.28) | 37<br>(0.79) |
| **MORE THAN 3 HOURS** | 44<br>(1.80) | 164<br>(3.59) | 8<br>(0.21) | 7<br>(0.38) | 2<br>(0.07) | 6<br>(0.29) | 1<br>(0.04) | 31<br>(0.84) | 5<br>(0.25) | 52<br>(1.40) | 2<br>(0.11) | 4<br>(0.14) | 3<br>(0.11) | 4<br>(0.30) | 5<br>(0.11) | 3<br>(0.08) |

Denote: Chi<sup>2</sup> significance: <sup>\*</sup> = *P*-value<0.05, <sup>\*\*</sup> = *P*-value<0.01, <sup>\*\*\*</sup> = *P*-value<0.001)

**Table 5:**
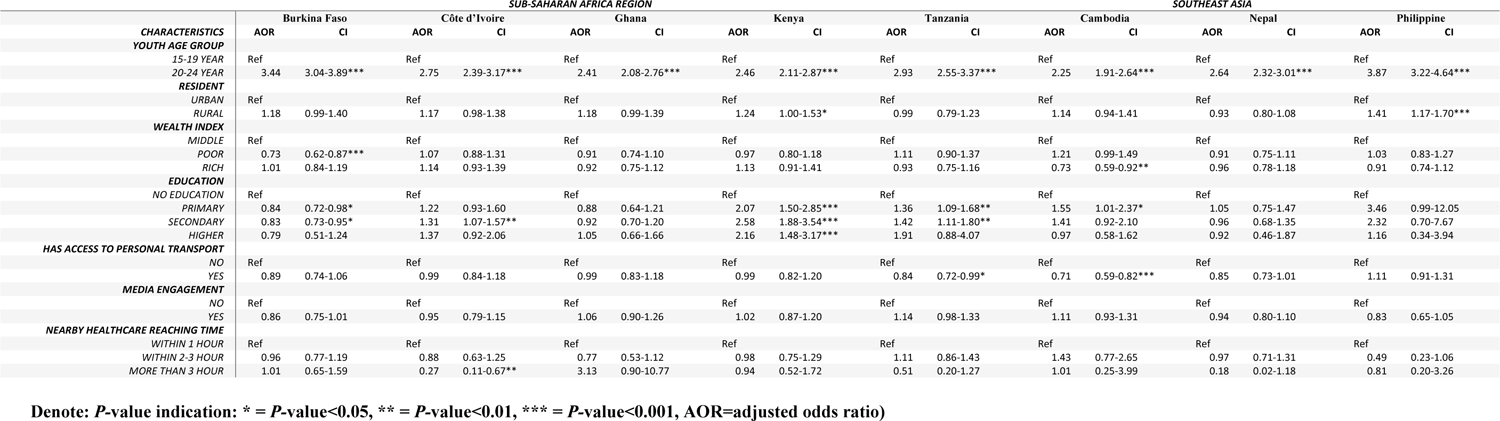
Influencing Factors of Healthcare Utilization among Female Youth Groups in Low and Lower-middle-income Countries.

On the other hand, multivariate analysis showed in Côte d’Ivoire that those who require more than 3 hours to reach nearby health facilities have a lower likelihood (AOR 0.27 CI:0.11-0.62) of healthcare utilization compared to those who have within 1-hour reaching time, and this is moderately statistically significant.

## Discussions

The comprehensive exploration of healthcare utilization among youth delves into intricate dynamics shaped by socio-demographic contexts, revealing variations across countries. The majority of participants are from the 15-19 age group, compared to the 20-24 age group, and most of them are from Burkina Faso. Additionally, most youths live in rural areas in the same country.

The study found a significant association across all regions of our study concerning urban-rural settings and the time required to reach nearby healthcare facilities. A notable trend has been observed: a higher percentage of individuals residing in urban regions reach healthcare facilities within one hour compared to those living in rural areas. This can be justified by Several interconnected factors, including urban areas typically receiving benefits from more developed infrastructure, including better road networks and transportation systems. The accessibility of healthcare allows urban residents to cover shorter distances and reach medical services more quickly. Also, the concentration of healthcare facilities tends to be higher in urban areas. This spatial distribution results in greater availability of medical services within close proximity in urban areas, facilitating prompt access. This may suggest that rural areas have fewer healthcare facilities, and individuals might need to travel more distances to reach the nearest center [34].

Across all the regions, there is a strong association between age group and healthcare utilization, and this is also reflected in multivariate analysis. In Burkina Faso, Côte d’Ivoire, Ghana, Kenya, Tanzania, Cambodia, Nepal, and Philippines, showed a higher likelihood of accessing healthcare services among the 20-24 years age group compared to the 15-19-year age group. This may suggest that the 20-24 years group may attribute to increased health awareness, emerging health needs, autonomy in decision-making, education, and improved access to resources during early adulthood, whereas the 15-19 years group may not have access to such resources and they may pose fear to do all this by themselves [35–37].

Throughout the study, we found education is one of the significant factors that influence healthcare utilization most. In Kenya, Tanzania, and Cambodia, individuals with primary education showed a higher likelihood of healthcare utilization compared to those without education. This suggests that having an education can benefit to receive more healthcare facilities and is supported by the belief that basic education helps develop health awareness and knowledge [38]. When individuals have received the education, they may acquire a foundation of health-related information, which empowers them to prioritize their well-being and seek appropriate healthcare services. Health literacy encompasses skills like accessing, understanding, evaluating, and applying health information. Through primary education this equips individuals with these skills enabling them to make decisions regarding their health [39]. In countries such as Côte d’Ivoire, Kenya, and Tanzania, individuals who have secondary education are more inclined to access healthcare services. This indicates that advanced education has a positive impact on their health-seeking behavior. In Africa, youth inequality and limited access to secondary education is very common issue for a long time. However, Innovations in Secondary Education (ISE) initiative addresses this, allocated US $35.5 million and doing twelve projects in several countries, including Côte d’Ivoire, Kenya, Malawi, Rwanda, Senegal, Tanzania, and Uganda. This could be a significant promoter of secondary education among youth in those regions, which could be a success of the program promoting secondary education and empowering youth with skills, thus making them dependent on receiving more healthcare utilization [40]. Moreover, in Kenya, youth who have received a higher education are more inclined to make use of healthcare services. Research indicates that individuals with higher education tend to have better health and longer lifespans compared to those with lower levels of education [41]. This suggests that advanced education equips individuals with thinking skills and a deeper comprehension of health issues, motivating them to seek out healthcare services. Despite this, Burkina Faso showed an inverse pattern in terms of both primary and secondary education. Burkina Faso showed that those with primary or secondary education have a lower likelihood of utilizing healthcare compared to those who are not educated. This is a very interesting finding. One possible reason may be that education may have improved the knowledge of individuals related to healthcare; thus, educated individuals take self-care rather than trying to go into healthcare rapidly compared to non-educated individuals [42]. Also, female youth may face restrictions to going outside rapidly, which could be due to their cultural beliefs, which can cause lower healthcare utilization [42].

Another key factor is residents, which showed an increased likelihood of healthcare utilization seen in rural individuals in Kenya and the Philippines compared to urban people. In Kenya’s context, remote regions may have a prevalence of infectious diseases [43]. Also, other conditions, including poor hygiene and suboptimal physical activity, may cause a burden among rural people compared to urban people, which may be a triggering factor for a higher likelihood of healthcare utilization.

On the other hand, the archipelagic nature of the Philippines presents obstacles to healthcare accessibility in isolated rural areas [44]. In rural settings, many may live in distinct mountain areas where healthcare access is not easy. Furthermore, there are more frequency of diseases and also, they don’t have access to other health facilities like pharmacy, shop where they can buy items for self-medication. As treatment may be available only in certain healthcare settings, which could be the possible reason that more female youth frequently rely on healthcare visits which are only available to them.

Moving onto the socio-economic context, in Burkina Faso, poor individuals show a lower likelihood of healthcare utilization compared to the middle class as they could be rooted in financial barriers. They may need help in affording healthcare, possibly due to the costs associated with accessing medical services. Since young women also do not earn enough, they may face financial issues, which may be the possible reason for less healthcare utilization [45,46]. In Cambodia, the rich are less likely to seek healthcare, as the wealthy can afford nutritional food and better accommodation which may cause lower prevalence of diseases among them. Moreover, low trust in the healthcare system, which has been affected by corruption, poor quality, and lack of accountability may hinder finding interest on receiving healthcare services in general [47]. Shifting focus to personal transport in Tanzania and Cambodia, the lower likelihood of healthcare utilization among those with access to personal transportation may opt for more distant to get healthcare facilities. This may create lack of interest on receiving healthcare services among youth female as it might not be easy for them to go frequently with their personal transport as they may need to rely on other members of their family for this [48].

The observed trend in Côte d’Ivoire, where individuals requiring more than 3 hours to reach nearby health facilities exhibit a lower likelihood of healthcare utilization compared to those within a 1-hour to reach. Côte d’Ivoire’s diverse terrain often poses obstacles to timely access to healthcare services. Moreover, lack of enough healthcare facilities can lead to such issues. People who have to travel distances may experience challenges with the transportation system, such as poorly maintained roads or a lack of convenient public transportation choices [49]. This detailed analysis highlights the relationship between socio-economic factors and geographical obstacles in influencing the way young people access healthcare services.

### Limitations

The study’s reliance on self-reported data which may introduces subjective biases. The temporal scope is limited to post-2020 data. Omission of countries due to insufficient data affects generalizability as representation of eight countries may not capture broader regional nuances. Uneven sample sizes across regions may introduce biases in precision and representativeness. Lack of exploration into participants’ cultural backgrounds misses insights into cultural influences on healthcare utilization patterns. These limitations suggest areas for future research to enhance understanding.

## Conclusions

This research shows that education plays a significant role in healthcare usage across the countries, with primary and secondary education positively influencing it. The connection between residence and the time it takes to get to healthcare facilities highlights difficulties among rural individuals when it comes to accessing services promptly. It’s fascinating that as young adults transition into adulthood between the ages of 20 and 24, they tend to become more conscious about their health, embrace their independence, and gain access to resources. However, Burkina Faso presents a contrast where educated individuals may prioritize self-care over seeking medical attention. Additionally, educated youth female may face barriers due to gender and cultural beliefs Burkina Faso, which result in lower healthcare utilization. Geography, diseases and limited accessibility, may further contributed to increased utilization of healthcare services in areas in Kenya and the Philippines. The situation in Côte d’Ivoire highlights an urgent need for intervention in the poorly maintained roads and convenient public transportation. Considering these findings, it is recommended that we focus on initiatives to enhance healthcare facilities in regions, overcome cultural and gender-related obstacles among female youth, and foster trust in health systems by promoting transparency and accountability. It is also important to focus on promoting female education and health literacy in different regions. Initiatives such as the UNESCO strategy on education for health and well-being should address education to empower female. Policymakers and healthcare professionals should work together to guarantee that health resources are distributed equitably and minimize the obstacles to healthcare facilities among youth.

## Abbreviations

ABBREVIATION: FULL FORM

AOR: Adjusted Odds Ratio

DHS: Demographic and Health Survey

EA: Enumeration Areas

ISE: Innovations in Secondary Education

SDG: Sustainable Development Goal

UNDP: United Nations Development Programme

## Data Availability

Data is available from the Demographic and Health Surveys (DHS) program. https://dhsprogram.com

## Acknowledgments

We thank the Demographics and Health Survey (DHS) for providing us with access to their survey data.

